# Large language models improve transferability of electronic health record-based predictions across countries and coding systems

**DOI:** 10.1101/2025.02.03.25321597

**Authors:** Matthias Kirchler, Matteo Ferro, Veronica Lorenzini, FinnGen, Christoph Lippert, Andrea Ganna

**Affiliations:** Hasso Plattner Institute, University of Potsdam, Digital Engineering Faculty, Potsdam, Germany; Hasso Plattner Institute for Digital Health, Icahn School of Medicine at Mount Sinai, NY, US; Institute for Molecular Medicine Finland (FIMM), HiLIFE, University of Helsinki, Helsinki, Finland; Analytic and Translational Genetics Unit, Department of Medicine, Massachusetts General Hospital, Boston, MA, USA

## Abstract

Variation in medical practices and reporting standards across healthcare systems limits the transferability of prediction models based on structured electronic health record (EHR) data. We introduce GRASP, a novel transformer-based architecture that enhances the generalizability of EHR-based prediction by embedding medical codes into a unified semantic space using a large language model. We applied GRASP to predict the onset of 21 diseases and all-cause mortality in over one million individuals from UK Biobank (UK), FinnGen (Finland) and Mount Sinai (USA), all harmonized to OMOP common data model. Trained on the UK Biobank and evaluated in FinnGen and Mount Sinai, GRASP achieved an average ΔC-index that was 83% and 35% higher than language-unaware models, respectively. GRASP also showed significantly higher correlations with polygenic risk scores for 62% of diseases. Notably, GRASP mantained robust performance even when datasets were not harmonized to the same data model, accurately predicting disease risk from ICD-10-CM codes without direct mappings to OMOP. GRASP enables accurate and transferable disease predictions across heterogeneous healthcare systems with minimal resource requirements.

## Introduction

Accurate disease risk estimation is important for guiding screening, preventative interventions, and early-stage treatments. Most disease prediction models used in clinical practice are based on a limited set of risk factors measured in prospective cohort studies^1–3^. However, the increasing availability of electronic health record (EHR) data, combined with advances in machine learning, has opened avenues for automated risk prediction directly from EHRs. For example, promising results have been shown in predicting diseases such as pancreatic cancer^4^ and cardiovascular conditions^5–7^.

Despite these opportunities, EHR data is inherently complex and lacks standardization across healthcare systems. Large providers with robust infrastructure can develop and deploy in-house models, but smaller healthcare often lack the necessary resources. Consequently, the ability to transfer predictive models across healthcare settings is crucial to democratizing access to EHR-based predictions.

A common approach to harmonizing health records involves mapping EHR data to a common data model (CDM), such as OMOP-CDM^8^, which uses standardized vocabularies like SNOMED or RxNorm. For example, harmonization efforts across EHR systems are envisioned by the European Health Data Space Act of the European Union. However, these efforts are resource-intensive, and even after standardization, EHR data remains heterogeneous due to factors such as local clinical practices, regulatory environments, and coding differences. This heterogeneity undermines the performance and generalizability of prediction models, limiting their applicability across diverse healthcare systems.

Researchers have shown that, despite the heterogeneous sources of medical codes vocabularies, it is possible to capture similarities between underlying medical concepts. This can be achieved by learning their latent representations using embedding-based approaches ^9–13^. Such embeddings, often derived from co-occurrence patterns, show promise but face generalization challenges when deployed externally. Rare medical concepts might occur insufficiently often in the training data to learn high-quality embeddings but may be highly relevant for accurate disease prediction. Other approaches derive fixed embeddings from external sources, such as clinical ontologies^14^. However, integrating explicit knowledge graphs often demands substantial effort to align disparate source of medical codes. A critical limitation of both ontology-based and co-occurrence-based embeddings is their dependency on the initial training vocabulary. Concepts absent during the embedding creation process cannot be incorporated during inference, restricting model adaptability in real-world deployment. This limitation becomes particularly pronounced as medical vocabularies evolve—many embeddings were trained exclusively on ICD-9 codes, rendering them obsolete for applications requiring ICD-10 or newer coding systems^11^. Some prior studies have explored direct text interpretation of medical concepts^15–17^, but they lack concrete applications for disease prediction across diverse healthcare systems.

We propose GRASP (Generalizable Risk Assessment with Semantic Projection), a novel deep learning approach designed to improve the transferability of EHR-based prediction models. Instead of relying solely on a common data model, GRASP maps medical concepts into a unified semantic space using a large language model (LLM). A downstream transformer network processes patient medical histories to predict disease risk, leveraging the LLM’s contextual understanding of medical concepts. GRASP bridges coding differences across EHR systems by leveraging learned similarities between medical concepts in the language space. For instance, GRASP successfully predicts disease risk even for medical codes absent from the training data.

GRASP is resource-efficient and can be deployed even in environments with low computational resources. The LLM is used exclusively to generate a lookup table of embeddings, eliminating the need to expose patient data directly to the model. This allows GRASP to operate in secure processing environments without internet connectivity. We applied GRASP to predict 21 diseases and all-cause mortality using EHR data from over one million individuals across three countries (United Kingdom, Finland, and the United States). Our results demonstrate that GRASP significantly improves model transferability across healthcare systems compared to state-of-the-art models like XGBoost^18^, even when data is harmonized to OMOP-CDM.

Notably, GRASP facilitates generalization across healthcare systems without need for harmonization of medical codes, leveraging semantic similarities to align otherwise distinct medical codes. The inductive bias of the LLM allows GRASP to achieve the same predictive performance as conventional models with far less training data, enhancing data efficiency. Furthermore, GRASP identifies individuals at higher genetic risk for diseases more accurately than language-unaware models, underscoring the value of integrating LLM-driven embeddings in disease risk prediction.

## Results

### GRASP architecture

Our goal is to predict the first occurrence of 22 health outcomes (21 diseases and all-cause mortality) over an average follow-up of 8 years using a patient’s OMOP-coded medical history (**Figure 1; Supplementary Tables 1, 2**).

**Figure 1.**
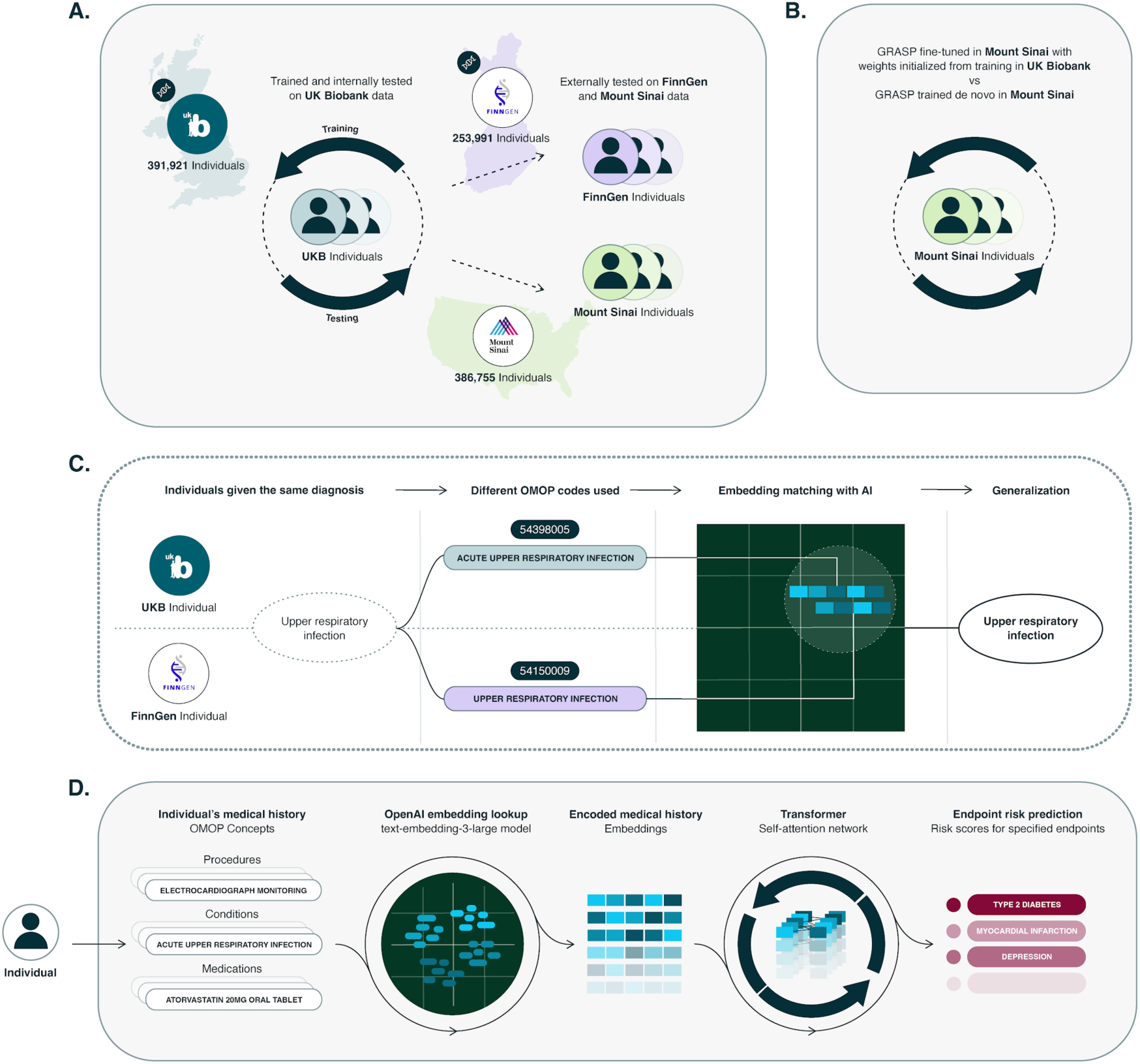
Overview of the study. **A.** model to jointly predict the first occurrence of 21 diseases and all-cause mortality is developed in UK Biobank and validated in FinnGen and Mount Sinai. **B.** We also evaluate the benefit of fine-tuning the model in Mount Sinai. **C.** Two patients have the same diagnosis but different OMOP codes, for example because of different coding practices. LLM embeddings can be used to match the two codes based on the corresponding natural-language name. **D.** GRASP maps the entire medical history of a patient using the text-embedding-3-large model from OpenAI and trains a transformer to predict the 22 outcomes. This allows to generate a score representing the disease risk for each outcome.

We begin by mapping all OMOP vocabulary concepts to semantic embeddings using a LLM (*OpenAI – text-embedding-3-large*). This step does not require individual-level data. The LLM processes the natural-language name or description of each concept (e.g., “Acute upper respiratory infection” instead of OMOP code 54398005) and generates a high-dimensional embedding. These embeddings form a lookup table that links every concept to its vector representation. A patient’s medical history is subsequently encoded by querying this lookup table, avoiding the need for repeated LLM evaluations during model inference.

Next, we introduce a multi-layer transformer neural network (**Methods**) that uses the encoded medical history and predicts the risk of developing each health outcomes^19^. Continuous variables (e.g., age) can be encoded with positional embeddings added onto the concepts. Unlike the LLM, the transformer architecture is lightweight, allowing efficient training and deployment. The network is trained jointly across all 22 endpoints using a Cox proportional hazards loss function.

The core advantage of GRASP stems from the LLM’s language understanding. For example, consider a scenario where the concept “High glucose level in blood” is prevalent in the source dataset, while “Hyperglycemia” is rare or absent. In the target dataset, the reverse holds true— only “Hyperglycemia” appears. The LLM embeds both concepts closely due to their semantic similarity. This allows the downstream transformer to generalize and recognize “High glucose level in blood” as near synonymous to “Hyperglycemia,” even if the latter was never encountered during training.

Crucially, this semantic alignment does not solely depend on surface-level similarities (e.g., spelling) but rather on the underlying meaning. By positioning semantically similar concepts closer and unrelated concepts farther apart, GRASP achieves greater data efficiency, enabling robust predictions even from small training sets. Additionally, GRASP facilitates zero-shot generalization to previously unseen concepts, enhancing model adaptability across datasets.

### Cohort characteristics and study design

We utilized three distinct datasets, including two large biobank-based studies: UK Biobank^20^ (UKB) from the United Kingdom (N=391,921), FinnGen^21^ from Finland (N=253,991), and one extensive EHR dataset from the Mount Sinai Health System in the United States (N=386,755). These datasets represent diverse populations, health systems and data collection methodologies.

Models were trained on UK Biobank data and evaluated with minimal adjustments on the FinnGen and Mount Sinai datasets. In UK Biobank, the baseline date was set to the first assessment center visit, while for FinnGen and Mount Sinai, a fixed date was uniformly applied across all individuals. To mitigate the risk of closely related conditions to the disease we aim to predict being used as predictors, we implemented a two-year washout period following the baseline date. Predictions were evaluated over the remaining follow-up period, which extended up to 11 years in UK Biobank, 6 years in Mount Sinai, and 10 years in FinnGen. We predicted the time-to-first event for all-cause mortality and 21 common diseases of significant public health relevance, as detailed in **Supplementary Table 1**. In UK Biobank, the condition with the highest incidence rate was knee osteoarthritis (4.3%), while inflammatory bowel disease had the lowest (0.41%).

Cohort characteristics and basic descriptors are provided in **Supplementary Table 2**. Age, sex, and OMOP-mapped concepts reflecting prior disease diagnoses, procedures, and drug prescriptions were used as predictors. On average, we observed 29 unique concepts per individual in UK Biobank, 39 in FinnGen, and 19 in Mount Sinai.

### GRASP improves performance and transferability across OMOP-mapped datasets

We first evaluated GRASP by comparing it to a baseline method that is language-unaware and uses randomized embeddings in place of those generated the LLM. This comparison allowed us to assess whether the semantic knowledge embedded in the LLM improves model performance. Additionally, we compared GRASP to XGBoost, a state-of-the-art approach for structured data^22^. Models were trained using 4-fold cross-validation on UK Biobank data for internal validation, and the resulting ensemble was evaluated on FinnGen and Mount Sinai data for external validation (**Methods**).

In the UK Biobank cross-validation test sets, both GRASP and the model using random embeddings consistently outperformed the age- and sex-based baseline (average ΔC-index of 0.068 and 0.080 for random embedding and GRASP model, respectively; **Figure 2, panel A; Supplementary Table 3**). However, the improvements with GRASP were larger, achieving an average 18% increase in ΔC-index across health outcomes compared to the language-unaware model. GRASP’s performance was comparable to that of XGBoost (**Supplementary Table 3**).

**Figure 2.**
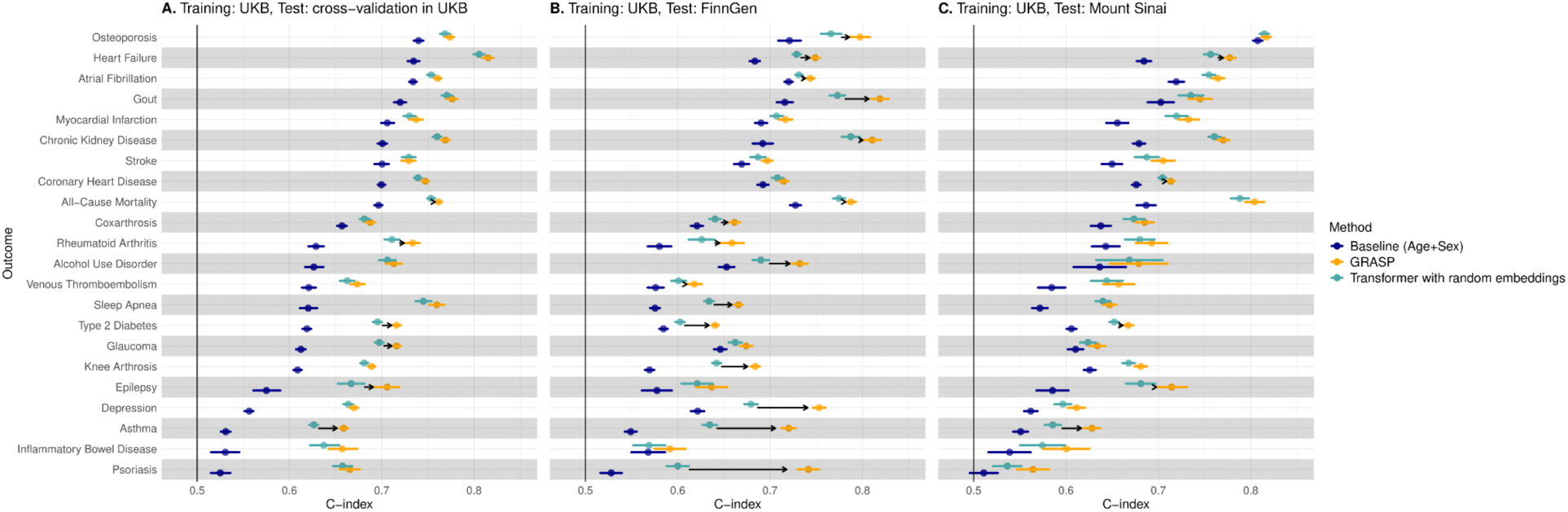
**GRASP model evaluation**. Performances (C-index) of a model with demographic (age and sex, blue) information only, random embeddings (light blue) and OpenAI embeddings (GRASP, orange) to predict the first occurrence of 22 outcomes. Models are trained in UK Biobank and tested in a held-out dataset from: **A.** UK Biobank, **B.** FinnGen and **C.** Mount Sinai. Horizontal lines represent 95% confidence intervals obtained via bootstrapping. Arrows are drawn if the difference between OpenAI embeddings and random embedding model performances are statistically significant at alpha=5%.

When applied to external datasets, GRASP demonstrated superior transferability. It outperformed random embeddings by 83% in FinnGen (average ΔC-index: 0.075 *vs.* 0.041; **Figure 2, panel B**) and by 35% in Mount Sinai (average ΔC-index: 0.062 *vs.* 0.046; **Figure 2, panel C**). Notably, GRASP also transferred more effectively than XGBoost in both external datasets (**Supplementary Tables 4 and 5**).

Next, we assessed whether adapting GRASP to the external dataset through fine-tuning could enhance its transferability. In the previous experiments, model weights remained frozen during external validation. Here, we fine-tuned the weights initially trained on UK Biobank data using a small, independent training set from Mount Sinai, separate from the test set. This approach allows GRASP to leverage the larger UK biobank dataset while adapting to the specific characteristics of the target data. For comparison, we evaluated GRASP trained solely on Mount Sinai data (without UK biobank pre-training) and XGBoost trained exclusively on Mount Sinai.

Fine-tuning improved GRASP’s performance relative to direct application, with an average C-index of 0.721 compared to 0.713 (**Supplementary Table 6**). Fine-tuned GRASP outperformed both XGBoost and the GRASP model trained exclusively on Mount Sinai data (**Supplementary Table 6**).

### GRASP transfers well across datasets mapped to different data models

So far, we have explored the transferability of GRASP on datasets mapped to a common data model (OMOP-CDM). Next, we investigated whether GRASP’s language-based embeddings could facilitate translation between different data models without requiring re-training. To assess this, we evaluated GRASP’s performance on Mount Sinai data under two conditions: (1) using only OMOP-mapped disease concepts, where the same data model was applied in both the training and testing datasets; and (2) using a different data model in the training set, where disease conditions were coded in the ICD-10-CM format. Notably, no explicit ontology mapping between SNOMED and ICD-10-CM was applied.

As expected, the highest performance was observed when both the training and test datasets were mapped to OMOP, resulting in an average ΔC-index of 0.056 compared to the age and sex-only baseline (**Figure 3**). However, even when evaluated using solely ICD-10-CM codes in Mount Sinai, GRASP demonstrated notable improvements, achieving an average ΔC-index of 0.036 over the baseline. This result is particularly striking given that no direct mapping between OMOP and ICD-10-CM was provided to GRASP. The model successfully inferred the relationships between the two coding systems by leveraging the semantic similarity of disease names across OMOP and ICD-10-CM concepts.

**Figure 3.**
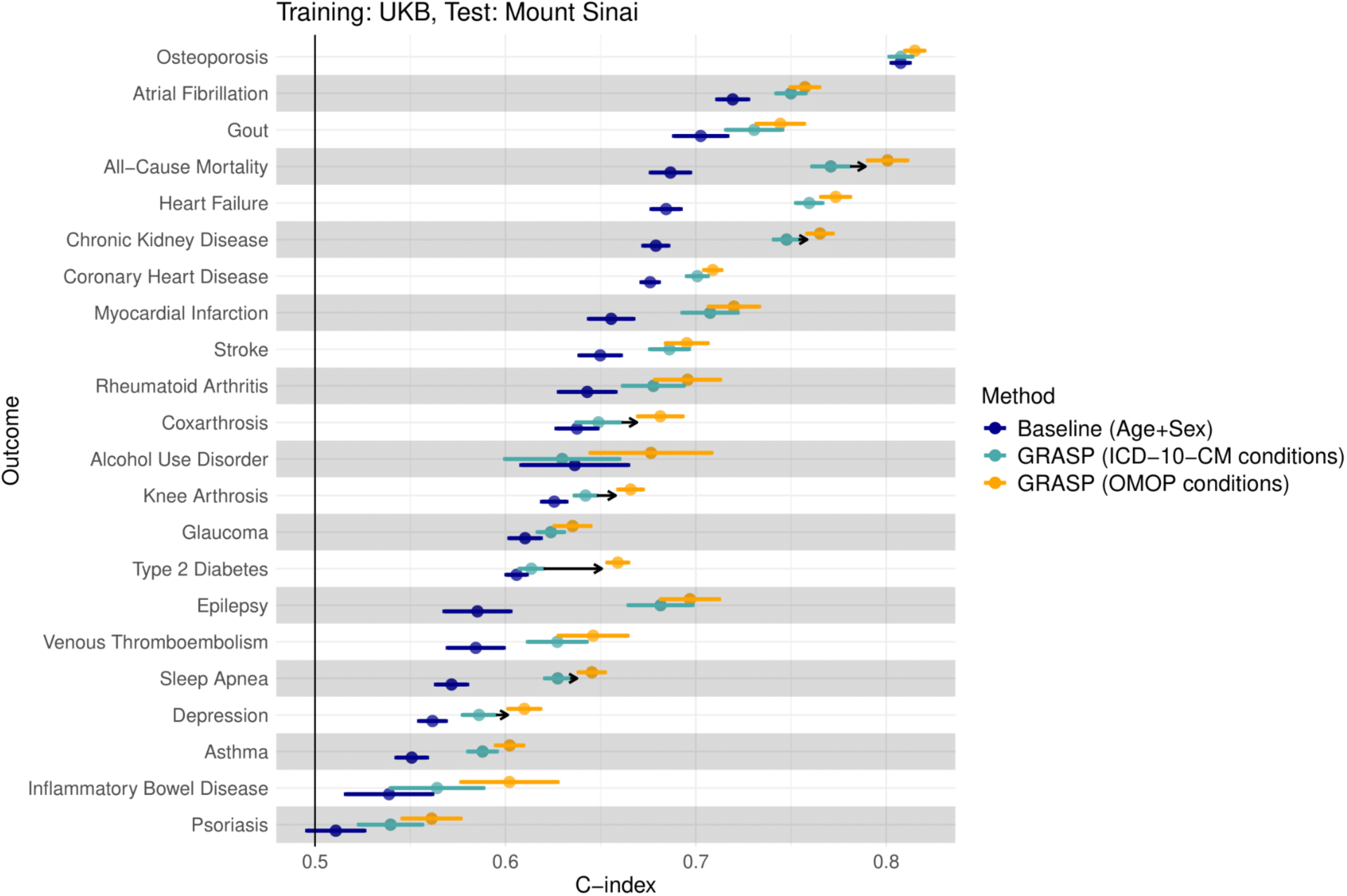
Transferability across datasets mapped to different data models. Models are trained in UK Biobank to jointly predict the first occurrence of 22 outcomes. The figure reports the performances in the Mount Sinai dataset of a model that just use age and sex (blue), GRASP model applied to a different data model (ICD-10-CM, light blue) or the same data model (OMOP-CDM, orange). Horizontal lines represent 95% confidence intervals obtained via bootstrapping. Arrows are drawn if the difference in performance between OMOP-based models and ICD-10-CM-based models are statistically significant at alpha=5%.

### GRASP improves training-efficiency with small sample sizes

We reasoned that GRASP introduces significant inductive bias by positioning similar concepts nearby and unrelated concepts far apart, which can result in more efficient data utilization with fewer individuals. To test this hypothesis, we re-train GRASP on subsets of 10,000 to 200,000 individuals in the UK Biobank and evaluate performances within UK Biobank via cross-validation and externally in FinnGen and Mount Sinai **(Methods)**.

We find that across the 22 health outcomes GRASP performances were significantly higher than for the same model with random embeddings and XGBoost. This effect was especially pronounced at very small sample sizes (**Figure 4** and **Supplementary Table 7**). For example, the average ΔC-index improvement against XGBoost was 0.1 at N=10,000 *vs* 0.01 at 200,000 in the cross-validation in UK Biobank. Unlike GRASP, XGBoost transferred poorly to Mount Sinai and FinnGen when trained on smaller sample sizes and significantly improved its performance only with larger sample sizes.

**Figure 4.**
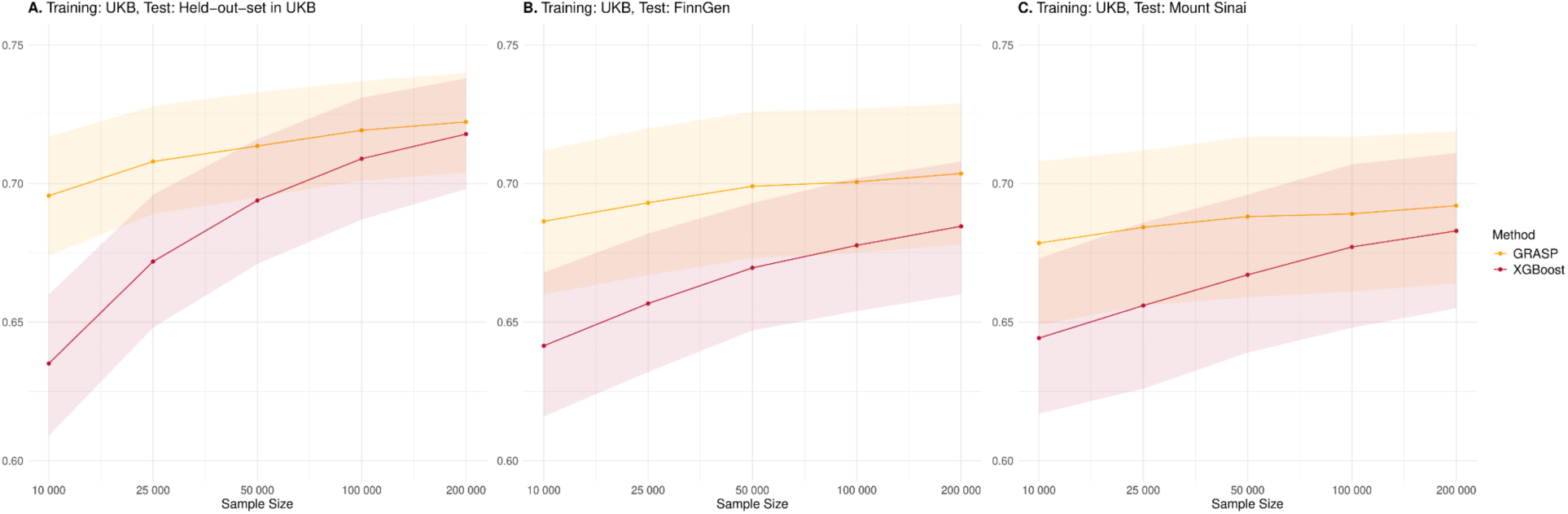
GRASP average prediction performances for different training sample sizes. Average prediction performances across 22 health outcomes as function of training sample size in UK Biobank. Comparison between GRASP (orange) and XGBoost (red). All models are trained in UK Biobank and evaluated in: **A.** the cross-validation test-sets of UK Biobank, **B.** FinnGen, and **C**. Mount Sinai. Shaded area represents 95% confidence intervals obtained via bootstrapping.

### Impact of concept-specific text on GRASP performance

We hypothesized that incorporating additional information about each concept/medical code could enhance model performance by providing a more comprehensive representation of its semantic and ontological context. To evaluate this hypothesis, we enriched the embedded text for each concept with supplementary data extracted directly from the OMOP ontology. This included hierarchical relationships such as concept ancestors and descendants, and attributes like associated morphologies and anatomical finding sites (**methods**). We found no substantial difference between using these enriched concept texts compared to the simpler embeddings using only the concept names (**Supplementary Table 8**), suggesting that just the name of the concept was sufficient to determine its similarly with other medical concepts.

### Understanding how GRASP generalize medical concepts

We wanted to better understand how semantic similarities between different OMOP codes can improve models’ performances and transferability. We use UMAP to provide a two-dimensional representation of the embedding representations of OMOP codes in UK Biobank, FinnGen and Mount Sinai focusing on depression, as an example (**Figure 5A**).

**Figure 5.**
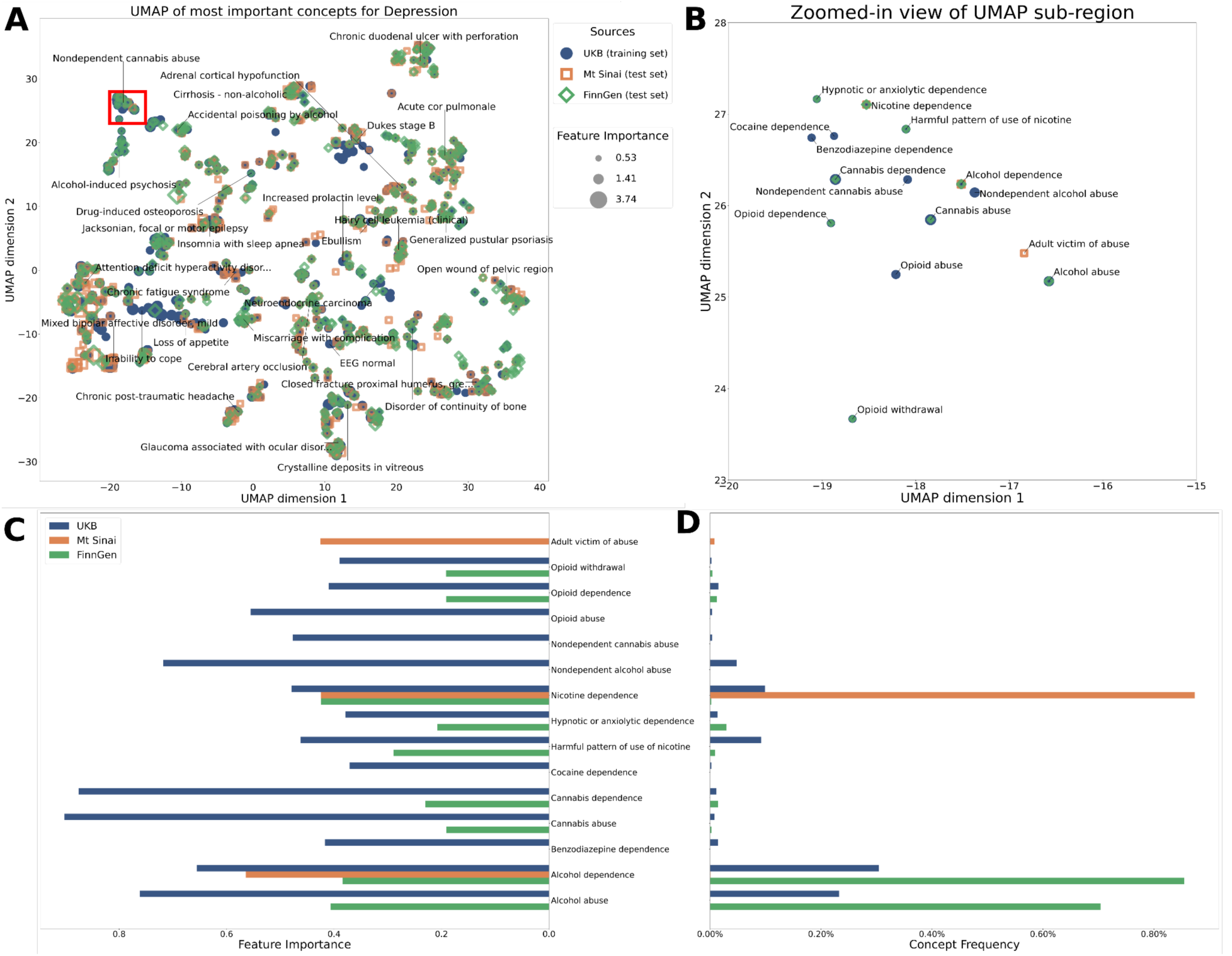
Representation of semantic embedding and feature importance across the three datasets for prediction of depression. **A.** UMAP of sematic embeddings for each OMOP concept contributing to prediction of depression in each study (identified by different shapes). The size of each shape indicates the feature importance of the concept. **B.** Zoom into the red box from panel A, identify a cluster of concepts related to substance abuse. **C.** Importance of each concept in the three studies (identified by different color). Not all concepts are present in all studies. **D.** Frequency of the concepts across the different studies.

We first show that similar concepts cluster together, even when they are present in only one of the three datasets. For example, we identified a cluster of concepts related to substance abuse. It is well-established that depression and substance abuse are closely linked^23^. Within this cluster, the concept of ‘opioid abuse,’ observed only in the UK Biobank, was closely positioned to ‘opioid dependence,’ which was also observed in FinnGen (**Figure 5B**). Similarly, ‘cocaine dependence,’ present only in the UK Biobank, clustered near other drug-related concepts. However, we also observed some clear misclassifications; for example, ‘adult victim of abuse’ clustered near drug abuse concepts, likely due to the shared term ‘abuse’ in their descriptions. Most concepts in the substance abuse cluster have similar importance in predicting depression (**Figure 5C**) despite large differences in frequency of concepts across biobanks (**Figure 5D**). These results highlight how the semantic embedding in GRASP overall helps overcoming differences in use and frequency of OMOP concepts across the three datasets.

### GRASP semantic embeddings result in stronger association with polygenic scores

We aimed to validate GRASP’s enhanced predictive capabilities through an orthogonal approach. Specifically, we hypothesized that GRASP’s risk estimates would more accurately identify individuals at higher genetic risk for diseases compared to models that do not incorporate language-based embeddings. Polygenic scores, which aggregate the effects of thousands of genetic variants, capture genetic risk and serve as an independent method to identify individuals at elevated disease risk. In FinnGen, we computed polygenic scores for 16 diseases following the approach described in Mars et al.^24^ and assessed their correlation with predictions from GRASP and a comparable model using random embeddings.

GRASP demonstrated significantly stronger correlations (p<0.05) with polygenic scores for 10 out of 16 diseases (**Figure 6**), indicating its superior ability to identify individuals with high genetic risk. These findings suggest that GRASP’s language-informed embeddings improve the model’s capacity to capture underlying disease susceptibility beyond what is achievable with language-unaware models.

**Figure 6.**
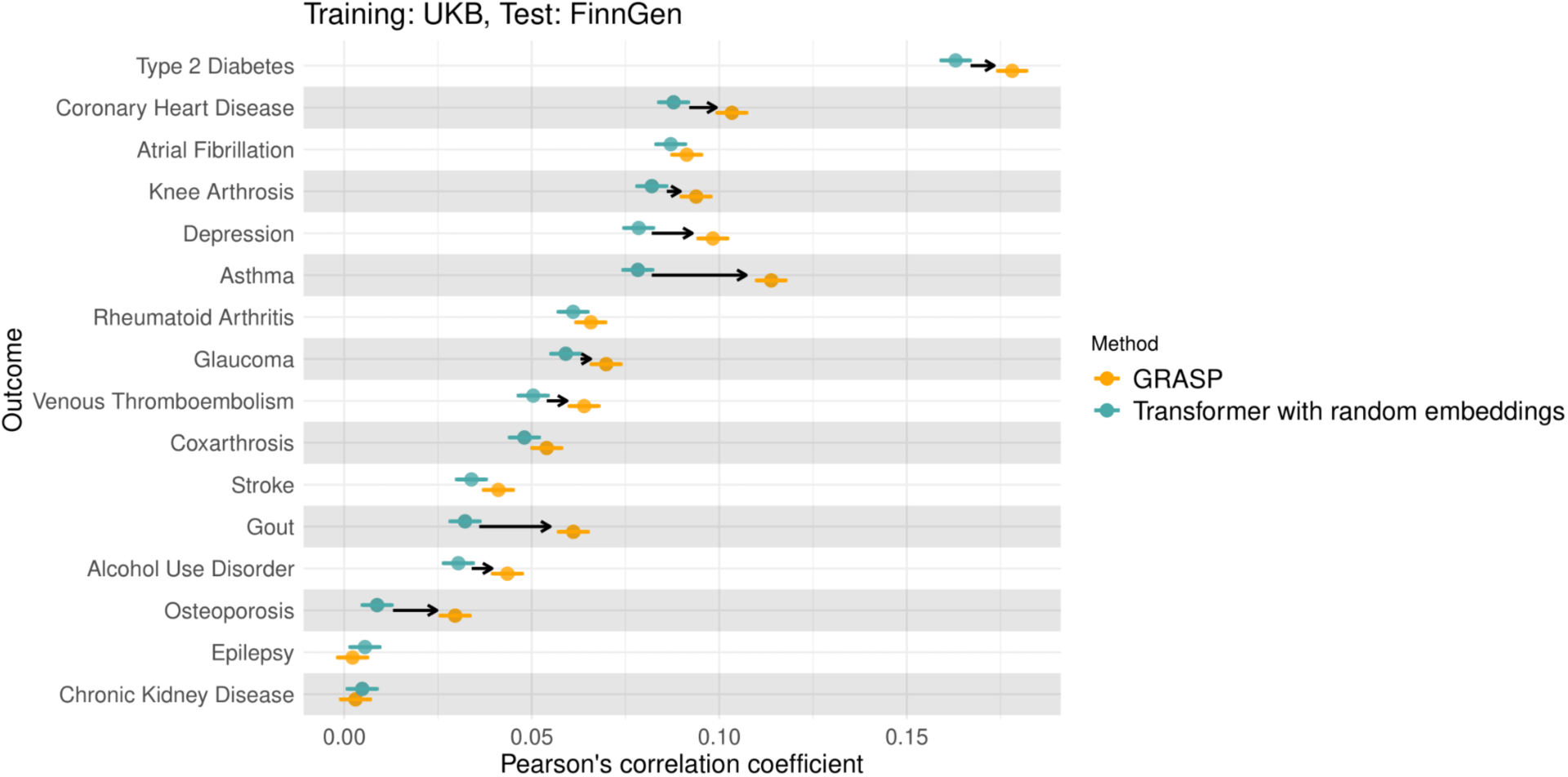
Correlation with polygenic score. Correlation between EHR-based risk score and polygenic score for 16 diseases in FinnGen, using GRASP with OpenAI embeddings (orange) and a baseline model with random embedding (light blue), both trained in UK Biobank. Bars represent 95% confidence interval. Arrows are drawn if difference between OpenAI embeddings and random embeddings are statistically significant at alpha=5%.

## Discussion

Harmonization of EHR data across healthcare systems through common data models (CDMs) is a valuable yet resource-intensive process. Initiatives like EHDEN have successfully promoted the adoption of the OMOP data model across European countries. However, the majority of EHR systems remain unmapped to OMOP, limiting interoperability. Even when datasets are fully harmonized to the same CDM, discrepancies persist due to variations in how medical codes are applied across healthcare systems.

Kather and colleagues recently argued that the reliance on standardized medical codes may be outdated in the era of LLMs, proposing that natural language should become the universal interface for healthcare^25^. While this vision holds promise, medical codes continue to serve critical roles beyond clinical practice, including healthcare management and billing. As such, it is unlikely that medical coding systems will be entirely replaced by natural language in the foreseeable future.

In this study, we present GRASP, a novel transformer-based architecture that predicts disease risk while addressing key challenges associated with the generalizability of medical codes. The core innovation of GRASP lies in embedding medical concepts into a unified semantic space using a large language model. This approach leverages the semantic similarity of medical terms, enhancing model transferability across different datasets and coding systems without the need for explicit manual mappings.

Our results demonstrate that GRASP improves the generalizability of EHR-based prediction models for 22 health outcomes across datasets from three countries. GRASP enhances model performance even when datasets are harmonized to the same CDM, and more importantly, facilitates transferability when datasets are not mapped to the same data model. By leveraging semantic language similarities rather than relying solely on direct mappings, GRASP bridges gaps in data interoperability, allowing for cross-system model deployment.

Johnson and colleagues recently introduced unified clinical vocabulary embeddings derived from a clinical knowledge graph, applying them to disease risk prediction in an Israeli healthcare system^14^. While this method offers an intriguing alternative, their embeddings couldn’t be map to all OMOP codes used in this study and lacked compatibility with ICD-10 codes. These limitations underscore the advantages of using embeddings derived from general-purpose LLMs, which provide broader coverage and are more easily integrated across diverse coding systems.

GRASP offers several key advantages. It is computationally efficient, fast to train, and can be deployed in secure, resource-limited environments without exposing patient-level data to external systems. Moreover, GRASP effectively utilizes small training sets by exploiting the inductive biases inherent in language models. Its ability to achieve zero-shot transferability across datasets with varying vocabularies or differing coding schemes highlights its potential to enhance data interoperability and improve predictive modeling across healthcare systems.

Nevertheless, several limitations remain. First, the current implementation of GRASP does not model the longitudinal sequence of medical codes, instead assuming that all codes are observed at a single point prior to baseline. Incorporating sequential architectures that capture the temporal progression of medical events—similar to approaches used in generative models like Delphi-2M^26^ —could enhance predictive performance. Additionally, integrating positional embeddings may enable GRASP to incorporate lab values, while future extensions could leverage online LLMs to analyze free-text clinical notes commonly found in EHR data. Second, GRASP’s performance has been evaluated using data from three high-income countries with advanced healthcare infrastructure. Its generalizability to underrepresented settings, including low- and middle-income countries, remains untested. Third, because the LLMs used in GRASP were not specifically trained on medical data, some embeddings may capture linguistic similarities that do not reflect true medical relationships. Furthermore, as with all models built on LLMs, GRASP may inherit biases from the language model’s training data, potentially reinforcing systemic inequalities or misrepresenting minority populations. Addressing these biases is critical to ensure equitable and safe deployment in real-world healthcare settings.

In conclusion, GRASP offers a potential solution to improve EHR-based disease predictions across diverse healthcare systems.

## Methods

### Datasets

#### UK Biobank

The UK Biobank^20^ is a large-scale biomedical database containing diverse health information from 500,000 middle-aged individuals recruited between 2006 and 2010 from across the UK. It includes extensive phenotypic information, genetic data, imaging, and electronic health records (EHR). We used EHR data mapped onto OMOP CDM by Regeneron and Odysseus Data Services and provided by the UK Biobank (field 20142). Data was originally sourced from hospital inpatient data, assessment center data, and partially from primary care data; however, primary care data is only available for ∼45% of the cohort and not necessarily complete. To reduce the likelihood of including individuals with incomplete data or those who may have transitioned between EHR systems, we restricted the cohort to participants with at least one recorded condition before and after the baseline date.

#### FinnGen

FinnGen (https://www.finngen.fi/en) launched in 2017, is a public-private research project, combining genome and digital healthcare data on about 500,000 Finns^21^. The nation-wide research project aims to provide novel medically and therapeutically relevant insight into human diseases. FinnGen is a pre-competitive partnership of Finnish biobanks and their background organizations (universities and university hospitals) and international pharmaceutical industry partners and Finnish biobank cooperative (FINBB). All FinnGen partners are listed here: https://www.finngen.fi/en/partners. List of FinnGen authors is provided in **Supplementary Table 9**.

#### Mount Sinai

The Mount Sinai Health System is a large network of hospitals and health-care providers in New York City. Longitudinal clinical data are recorded and mapped to the OMOP CDM in the Mount Sinai Data Warehouse (MSDW), and data are made available to researchers via the AI-ready Mount Sinai (AIR·MS) platform. The MSDW contains records for more than 11 million patients across more than 87 million patient encounters and is updated regularly. It provides longitudinal data on diagnoses, lab results, prescriptions, hospitalizations, and procedures. Due to the urban location of the hospital system, this dataset consists of a highly diverse population, representing multiple ethnicities and socioeconomic groups. To reduce the likelihood of including individuals with incomplete data or those who may have transitioned between EHR systems, we restricted the cohort to participants with at least one recorded condition before and after the baseline date.

### Endpoint definitions

Even though all three datasets are mapped to the OMOP CDM, they used partially different mappings. This makes creating homogeneously defined, clinically meaningful endpoints challenging. We decided to utilize pre-defined, harmonized endpoints from the FinnGen project. These were originally defined using ICD revisions 8, 9, and 10, as well as additional information from ICD-O-3, procedure codes, drug reimbursement codes and ATC codes. For FinnGen individuals, all endpoints were already precomputed. For Mount Sinai data, we used the original ICD-10-CM codes and defined endpoints using the ICD-10 FinnGen definitions.

In UK Biobank we had less detailed access to ICD-mapped codes with significant discrepancy between OMOP-mappings and ICD mappings which required a more complex workaround. In particular, we used ICD-10 codes from FinnGen as a starting point and mapped those to SNOMED conditions using the OMOP ontology (“Non-standard to Standard map”/”maps to” relationship, using both ICD-10 and ICD-10-CM). Unfortunately, this mapping was error prone and we manually filtered incorrectly mapped concepts (e.g., OMOP concept “Periodontal disease” (OMOP ID 134398) mapped to ICD-10-CM “Type 2 diabetes mellitus with periodontal disease” (ICD-10-CM code E11.630), which is a subcategory of “Type 2 diabetes mellitus” (E11)). To additionally reduce the amount of endpoint leakage, for each endpoint we manually inspected the concepts with highest occurrence discrepancy between cases and controls and added these concepts if needed. The final endpoint was then defined from this list of OMOP concepts. Despite these quality controls, a small amount of endpoint leakage from OMOP concepts is to be expected across all three datasets, slightly inflating model performance for all evaluated methods. Only for the endpoint “Type 2 diabetes” we additionally created a list of concepts to exclude individuals with such an occurrence; we excluded individuals if T2D could not be distinguished from other types of diabetes (e.g., the concept “Autonomic neuropathy due to diabetes mellitus”), or if the individual shows another type of diabetes (e.g., “Type 1 diabetes mellitus uncontrolled”).

### Main experiments setup: Training in UK Biobank

We train all EHR models on UK Biobank data using 4-fold cross-validation, using a similar evaluation strategy to Steinfeldt et al.^27^. We created four train/test splits with disjoint test sets where approximately 75% of the data are training and 25% of the data are test sets; we split folds along assessment centers to reduce bias and data leakage due to regional stratification and assessment personnel & equipment. Within each training set we set apart 10% (relative) for validation and hyperparameter selection. All model setups use identical data splits.

Reported evaluations on UK biobank data denote the overall c-index when aggregating all test-split predictions from all four models (for all model setups).

### Main experiments setup: Application in external datasets

In the external datasets we have four independent models (one for each train/test split in UK Biobank) and use those as an ensemble to generate four risk scores for each individual and endpoint. We average all four scores to get an overall risk score and feed this risk score together with age and sex into a simple Cox-PH model to adjust the model to the target setting, using 20% of the target data. We don’t adjust the risk-model itself, except if explicitly noted.

### Grasp architecture and design

The GRASP architecture consists of three phases (Fig 1**, Panel D**). First, we map the full EHR vocabulary onto semantic embeddings using a large language model. Second, we use these embeddings as input to a multi-layer transformer neural network to predict risk scores for all target endpoints jointly. Finally, for each endpoint, we tune a linear Cox Proportional Hazards model on only age, sex, and the EHR-based risk score.

#### Step 1: LLM embedding setup

Implementations of OMOP all depend on a common set of standard vocabularies, such as SNOMED for conditions and procedures or RxNorm for drug prescriptions. While datasets can use additional non-standard vocabularies or custom concepts, the overwhelming majority of are either already available in the standard vocabularies or can be mapped using non-standard to standard mappings provided by OMOP (e.g., from ICD-10-CM to SNOMED). In the first step of the GRASP architecture, we embed all open OMOP standard vocabularies into a semantic embedding space using OpenAI’s state-of-the-art embedding large language model (“text-embedding-3-large”). For this, we only take the concept name, e.g., “Hyperglycemia” for the concept with OMOP Concept ID 4214376 and ignore other metadata (see **Suppl Table 8** for experiments with additional contextual information). Concept names can have varying length, with conditions and procedures often relatively short (e.g., “Essential hypertension”, “Cough”, or “Radiologic examination of knee”) and concept names for drug exposures sometimes including a list of active components and brand names (e.g., “floxacillin 250 MG Oral Capsule” or “60 ACTUAT fluticasone propionate 0.25 MG/ACTUAT / salmeterol 0.05 MG/ACTUAT Dry Powder Inhaler [Wixela]”).

We once created a large lookup table from all available concepts. Notably, this did not require the use of any dataset-specific data at all: we mapped standard and valid conditions (n= 173,526), procedures (n=254,010), and drugs (n=2,007,406) from the freely available standard vocabularies as provided by OHDSI. This excluded, however, the non-free procedure vocabulary CPT-4 commonly used in the US, including the Mount Sinai dataset, which we mapped back to SNOMED procedures using the OMOP “CPT-4 to SNOMED equivalent” relationship. After creation of the lookup table our approach did not require any more access to a large language model and did not expose any patient data directly or indirectly to third parties.

In addition to the conditions, procedures, and drug exposures we also created embeddings for sex (with text “Biological sex is female” or “Biological sex is male”) and age (with text “Age at baseline”, see next section for details on age encoding).

The resulting embedding has a dimensionality of 3072 and is normalized to unit Euclidean norm. Semantically similar concepts are positioned close together even if they don’t coincide on a character level since^28^.

#### Step 2: Model architecture and training

In traditional machine learning settings, an individual’s medical history would be required to be encoded as a single vector of fixed length, with each dimension likely corresponding to a single concept. By contrast, in our setup, an individual medical history *h_i_* is represented as the variable-size collection of encountered OMOP concepts, mapped to their respective

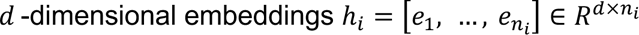

To encode an individual’s age, we incorporate a sinusoidal positional embedding^19^ into the “age at baseline” embedding. For other concepts we omit associated quantitative information, such as time of occurrence or sequence order, though these could also be included using positional embeddings. We retain all occurrences of each concept rather than keeping only a single occurrence of each unique concept.

Our primary model is based on a standard transformer encoder architecture^19^ without causal masking. For a given individual *i*, the transformer linearly maps all input embeddings [*e*_1_ …, *e_n_i__*] to 256-dimensional tokens and adds a learnable class token, creating the input to the first transformer layer, 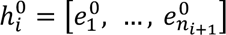.

Each transformer layer transforms input 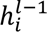 to output 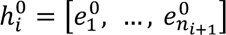 with identical dimensions.

Each transformer layer applies two main components: (1) a multi-head attention (MHA) mechanism with scaled dot-product attention, and (2) a fully-connected multi-layer perceptron (MLP). Both components are integrated with layer-normalization^29^ and a residual connection. The multi-head attention layer combines several smaller attention blocks (“heads”) for computational efficiency. Intuitively, these attention blocks dynamically relate each input token 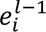 to all other input tokens in the same layer 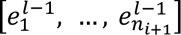, to determine which tokens (i.e., concepts) to focus on. The MLP block then non-linearly transforms the output of the MHA to enable the model to learn more complex features. In our case, the MLP consists of two linear layers with intermediate dimension 1,024 and GELU nonlinearity. After processing through all L layers, we discard the token-specific embedding outputs 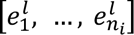 and use only the class token embedding for the final prediction. A linear layer on top of the class token generates risk scores for each endpoint.

We used L=4 layers and multi-head attention with 8 attention heads.

In contrast to more traditional machine learning models, a transformer network can process inputs with varying length *n_i_*. During training, however, we randomly sample n = 64 concepts per individual to enable faster training with homogeneously sized batches. If less than 64 concepts are available for an individual, we pad the available concepts with 0-valued empty concepts. During inference, we always include all concepts for an individual without padding.

We use mini-batch stochastic gradient descent with the AdamW optimizer^30^ to train each model for all 22 endpoints jointly with a batch-wise Cox log-partial likelihood from the Cox Proportional Hazards model^31,32^. We average the loss over all endpoints. If the loss for a single endpoint is not well-defined, we drop that endpoint’s loss for that batch, implicitly oversampling uncensored individuals. After hyperparameter selection in the UK Biobank cross-validation setup, we train all models with a batch-size of 512 for 8 epochs using a base learning rate of 0.001 with linear learning rate warmup for 2 epochs and cosine decay afterwards.

#### Step 3: Model application

The main model is trained on all endpoints and all individuals jointly on UK Biobank data, but this includes individuals with endpoint occurrences before baseline. For each endpoint, we fit an additional (linear) Cox-PH model using the lifelines Python library v0.27.8. This model takes as input age, sex, and the EHR-based risk score and performs time-to-event prediction with proper exclusion of prior events of the same endpoint. We perform this final adjustment on the same training sets in the UK Biobank as the main model training. In the two external datasets (FinnGen and Mount Sinai), we perform this tuning on 20% of the full dataset and evaluate on the remaining data.

### Baseline Methods

We compare GRASP against three baselines: (1) we compare against a model with identical architectural setup but with (fixed) randomized embeddings, to disentangle how much of GRASP’s performance is due to the use of embeddings versus architectural design; (2) we compare against gradient boosting trees (XGBoost) to compare against a powerful state-of-the-art prediction model; and (3) we compare against a simple Cox-PH model using only age and sex.

#### Random Embeddings

We use identical setup and architecture in this setting. The only difference is that embeddings are drawn at random from a Gaussian distribution with Euclidean normalization. While GRASP can adapt to unseen concepts in the target dataset with zero-shot adaptation, this is not possible for random embeddings, and we only use concepts that have been seen during training. We performed identical hyperparameter search as for GRASP but found that both models are stable with respect to architectural details such as number of layers, embedding dimensions, and optimization parameters, and used identical settings.

#### Gradient Boosting (XGBoost)

As a more powerful baseline model, we used gradient boosting algorithm^18^. We encode an individual’s medical history as a 0-1-coded (or count-coded) p-dimensional vector, where each dimension denotes the occurrence or absence of the associated concept. In contrast to GRASP, this model can only handle fixed-size inputs and can only utilize concepts seen during training. To enable better generalizability of the model, we used OMOP’s ontology to also activate ancestor features of each concept. XGBoost implements survival prediction with an accelerated failure time model instead of the Cox proportional hazards model. We performed hyperparameter search over the minimum number of occurrences required to include a concept (set to include all), whether to include concept occurrence counts or binarized indicators (set to binarized), learning rate (set to 0.1), max depth per tree (set to 2), number of boosting rounds (set to 500), scale of the AFT loss distribution (set to 3.0), and the number of ancestors to turn on in the ontology (set to 5) in the UK Biobank cross-validation.

Since we trained boosting models already per endpoint instead of jointly for all endpoints, in UK Biobank evaluations we used the predictions directly without the three-variable Cox-PH model on top. For adjusting predictions in the external datasets, we used the same setup as for GRASP, namely, first applying the boosting model to get a risk score that we then used together with age and sex as input to a Cox-PH model trained on 20% of the full dataset and evaluate on the remaining data.

### OMOP-to-ICD experiment

To assess the models’ transferability across coding schemes, we used the same model trained on UK Biobank data from all used OMOP tables (conditions, procedures, drugs, and age and sex), but evaluated them on either only the condition table in Mount Sinai, or on condition data coded in the original ICD-10-CM vocabulary (both together with age and sex).

Instead of running the full ensemble of all four models, we only use a single model trained on UK Biobank data. For the OMOP condition-only evaluation, we predict EHR-based risk scores identically to the previous setting but discarding procedure and drug exposure data for better comparability.

We created embeddings for ICD-10-CM data analogously to OMOP-coded data, by mapping each possible concept name directly with the same large language model to a semantic embedding. As data on conditions in the Mount Sinai data warehouse were originally recorded in ICD-10-CM format, we could use the original concept codes instead of mapping OMOP concepts to ICD codes via the ontology.

The remaining setup was identical to the main experiments.

### Small-n experiment

To simulate smaller sample sizes, we subset the UK Biobank training data to 10,000, 25,000, 50,0000, 100,000, and 200,000 individuals, respectively. We use the same train-test cross-validation splits from the main experimental setup. However, we keep the validation sets identical across all settings (10% of the original training split) and only subset the remaining training data to the respective dataset size to ensure comparability across splits. The test splits remain the same across all settings.

Like in the main experiments we train individual models per cross-validation split and apply all four models in the external datasets for an averaged risk score per individual.

### Ontology-enriched embeddings experiment

We designed more detailed texts for each possible concept to investigate if additional context or information beyond the concept name would improve embedding performance and therefore model performance. We used the OMOP ontology to create this enriched concept text with synonyms and related concepts. For a given concept, we collected all available synonyms and all pairwise relationships with the concept as the first of the two. We created a new string from this information in the form “{concept_name}. Synonym: {synonym1}. Synonym: {synonym2}. […] {relationship_name}: {other_concept} […]”. For drugs we limited the maximum number of synonyms to 10. To keep string lengths manageable, we also excluded approximately 100 relationship types, mostly consisting of mappings between different drug coding systems. The resulting string, for example for concept 43530605, is “Pulmonary embolism with pulmonary infarction. Synonym: Pulmonary embolism with pulmonary infarction (disorder). Synonym: Pulmonary embolism with infarction. Is a: Pulmonary infarction. Is a: Injury of artery. Has finding site: Structure of artery of pulmonary circulation. Is a: Pulmonary embolism. Has associated morphology: Embolus. Has finding site: Lung structure. Has associated morphology: Infarct.”

We replaced the original embeddings with these enriched embeddings but kept all other settings identical to the main setup.

### Fine-tuning experiment

For the fine-tuning experiment on Mount Sinai data, we used the same train and test splits as for the main evaluation where we only tuned the Cox-PH model on top of the frozen model trained on UK Biobank.

Instead of evaluating all four ensemble models as in the main setting, we only trained and evaluated a single model per setting. For hyperparameter selection we performed a 75%/25% train-validation split on the 20% of the data separated for training. We trained three separate models: (1) re-trainining GRASP from scratch; (2) fine-tuning GRASP with weights initialized from training in UK Biobank; and (3) training an XGBoost model from scratch.

For both the fine-tuning and the retraining from scratch we used identical optimization settings, training for 8 epochs with a batch size of 512, and randomly sampling 64 tokens per individual. For fine-tuning we did not freeze any layers as preliminary results did not indicate any improved performance.

Due to the different set of available concepts between UK Biobank and Mount Sinai, there is no straightforward way to fine-tune boosting models between datasets. Instead, we train the XGBoost model only from scratch. We used the same train-validation splits as for the GRASP models and performed a hyperparameter selection over the loss distribution scale, while keeping other training parameters identical to the training in UK Biobank.

We evaluated all models on the same large test set of 80% of the Mount Sinai data as the main experiments were performed on.

### Explainability

We require feature importances for each individual concept instead of per input dimension as is common in traditional machine learning settings or simpler fully connected neural networks. We derived concept-level feature importances for a single individual and endpoint by evaluating the model’s predicted risk score after occluding (i.e., setting to 0) the full embedding for each available input concept for that individual and subtracting it from the originally predicted risk score. We only used unique occurrences of concepts per individual. If an individual has k unique concepts, computation of this score would require k + 1 model evaluations. If a concept increases the individual’s risk for the endpoint, the feature importance score will be positive, otherwise negative or close to 0. To get overall feature importance scores for a dataset, we created these occlusion scores for all individuals in the dataset and averaged them over all individuals with this concept. Hence, these feature importance scores are dependent on the endpoint and on the dataset they were evaluated on; if a concept is not available in a dataset there is no associated feature importance score. Due to computational constraints, we only evaluated scores for a single model (instead of all four ensemble models). To better contrast the movement of concepts, in UK Biobank we evaluated the scores on the training set, in the external datasets on all individuals.

For the plot in **Figure 5**, we filtered all available condition concepts for minimum count of 5 occurrences in at least one of the three datasets and retained only the top 700 concepts according to their feature importance (using maximum over all three datasets) and applied a UMAP dimensionality reduction.

### Polygenic scores

Polygenic scores used were previously calculated by Mars et al ^24^ in FinnGen and are available at PGS Catalog (https://www.pgscatalog.org/, publication ID PGP000364)

## Data Availability

The code for the project is available at https://github.com/mkirchler/grasp. The individual-level data in these studies is protected for data privacy, access is regulated through the biobanks. The Finnish biobank data can be accessed through the Fingenious services (https://site.fingenious.fi/en/) managed by FINBB. UK Biobank data are available through a procedure described at http://www.ukbiobank.ac.uk. Mount Sinai EHR data can be accessed via use agreement with researchers at Mount Sinai.

## Acknowledgements

This research was conducted using the UK Biobank Resource under Application Number 77717. It was supported by the Office of Research Infrastructure of the National Institutes of Health (award number S10OD026880), the AI-Ready Mount Sinai (AIR·MS) research platform (developed through collaboration between the Hasso Plattner Institute for Digital Health at Mount Sinai and Data4Life), and the computational and data resources provided by the Icahn School of Medicine at Mount Sinai, supported by the Clinical and Translational Science Awards (CTSA) grant UL1TR004419 from the National Center for Advancing Translational Sciences. Additionally, the research was funded by the European Commission in the Horizon 2020 project INTERVENE (Grant agreement ID: 101016775). A.G. received funding from the European Research Council under the Horizon 2020 research and innovation programme (grant number 945733) and from the Academy of Finland fellowship grant no. 323116.

## Data and code availability

The code for the project is available at https://github.com/mkirchler/grasp. The individual-level data in these studies is protected for data privacy, access is regulated through the biobanks. The Finnish biobank data can be accessed through the Fingenious® services (https://site.fingenious.fi/en/) managed by FINBB. UK Biobank data are available through a procedure described at http://www.ukbiobank.ac.uk. Mount Sinai EHR data can be accessed via use agreement with researchers at Mount Sinai.

